# Estimating the harms from smoking and second-hand smoke exposure in social housing: a modelling study

**DOI:** 10.1101/2025.09.22.25336399

**Authors:** Samantha Howe, Tim Wilson, Kylie Morphett, Kate Mason, Germaine Lai, Vaughan W. Rees, Driss Ait Ouakrim

**Affiliations:** School of Population and Global Health, University of Melbourne; School of Public Health, The University of Queensland; NHMRC Centre of Research Excellence on the Tobacco Endgame; Harvard T.H. Chan School of Public Health

## Abstract

**Background:** People living in social housing in Australia have higher prevalence of daily smoking and greater exposure to second-hand smoke (SHS), particularly in multi-unit housing where smoke can drift between dwellings. The health impact of this exposure has not been quantified for this population.

**Methods:** We developed a Monte-Carlo matrix model of household SHS exposure, including smoke drift between dwellings in multi-unit housing, and linked it to the *SHINE Tobacco* simulation platform. The model projected health outcomes for the population living in social housing in the state of Victoria, Australia, over 20 years, comparing scenarios that eradicated SHS exposure and/or smoking with a business-as-usual (BAU) scenario of constant smoking prevalence. Outcomes were health-adjusted life years (HALYs) gained, and premature deaths averted across 31 smoking-attributable and eight SHS-attributable diseases.

**Findings:** Eradicating SHS exposure within the social housing population of Victoria in 2025 could result in 5,350 HALYs (95% uncertainty interval [UI] 4,670-6,120) gained, and 600 premature deaths (95% UI 500-700) averted over 20 years. In multi-unit housing, about half of the SHS-related health gain was attributable to eliminating smoke drift between units. Overall, SHS eradication accounted for approximately 27% of the total health gain achievable if tobacco smoking were fully eradicated in this setting.

**Interpretation:** Reducing SHS exposure in social housing would deliver substantial health benefits, with a large share resulting from preventing smoke drift in multi-unit housing. Better data on population dynamics and smoke infiltration would strengthen estimates and support policy design.

**Funding:** This research was funded by a seed funding grant awarded by the NHMRC Centre of Research Excellence on Achieving the Tobacco Endgame (NHMRC, GNT1198301)

## Introduction

Residents of social housing (housing provided or subsidised by public agencies) are at higher risk of experiencing tobacco-related harms. In Australia, approximately 34% of adults (age 15+) in social housing smoke daily, more than three times the national average.^1^ Half of people who smoke in social housing report doing so in their homes, compared to 15% among the broader smoking population.^2^ Consequently, residents of social housing, including those who do not smoke, are also more likely to experience greater second-hand smoke exposure in the home. Second-hand smoke contains a mix of potent carcinogens and other toxic chemicals, and fine particles known to increase the risk of cancer, as well as cardiovascular and respiratory diseases.^3^ Exposure is amplified in multi-unit housing due to smoke incursion or ‘drift’ from neighbouring dwellings. Around 40% of Australian social housing consists of multi-unit housing, compared to 16% in other tenure types.^4,5^

The Australian National Tobacco Strategy 2023–2030 (NTS) identifies reducing tobacco exposure in social housing and multi-unit housing as priority areas of action.^6^ Currently, no nationwide smoke-free regulations cover private dwellings in social housing or in multi-unit housing more broadly. Some jurisdictions impose partial restrictions, such as Queensland’s ban on smoking in common areas of multi-unit housing buildings.^7^ Internationally, stronger approaches have been implemented. For example, the United States introduced a federal ban in 2018 on smoking inside public housing buildings and within a set distance from entrances.^8^ While the evaluation of this policy shows mixed results on air quality and smoking cessation outcomes^9^, it signals growing recognition of the need to address second-hand smoke exposure in high-density, low-income housing.

Despite these policy developments and the known disparity in tobacco smoke exposure, the specific health burden attributable to smoking and second-hand smoke exposure in Australian social housing remains unquantified. Existing burden of disease studies, such as the Global Burden of Disease Study (GBD), and Australian Burden of Disease Study, rely on self-reported second-hand smoke exposure at home or work.^10,11^ While suitable for broad population analyses, these measures fail to capture higher-intensity exposures, due for example to smoke drift between units.

High quality evidence is needed to inform effective policies to protect social housing residents from second-hand smoke. To address this gap, we developed a simulation model integrating housing density, smoking prevalence, and spatial patterns of smoke drift to estimate health impacts from both direct smoking and second-hand smoke exposure. Using the state of Victoria as a case study, we quantified the potential health gains under hypothetical tobacco control scenarios, including the complete eradication of smoking and second-hand smoke exposure.

Our objectives are twofold: 1. to model the potential impact of tobacco-control interventions in social housing specifically, and 2. to account for differences in second-hand smoke exposure between multi-unit housing and stand-alone dwellings.

## Methods

### Overview

We used *SHINE Tobacco*, an established tobacco policy simulation model^12,13^ to forecast smoking prevalence and associated health outcomes among the social housing population in Victoria, Australia (population ∼ 7 million people, with social housing residents comprising 2·2%).^4,14^ Characteristics of this population are provided in Table 1.

**Table 1.**
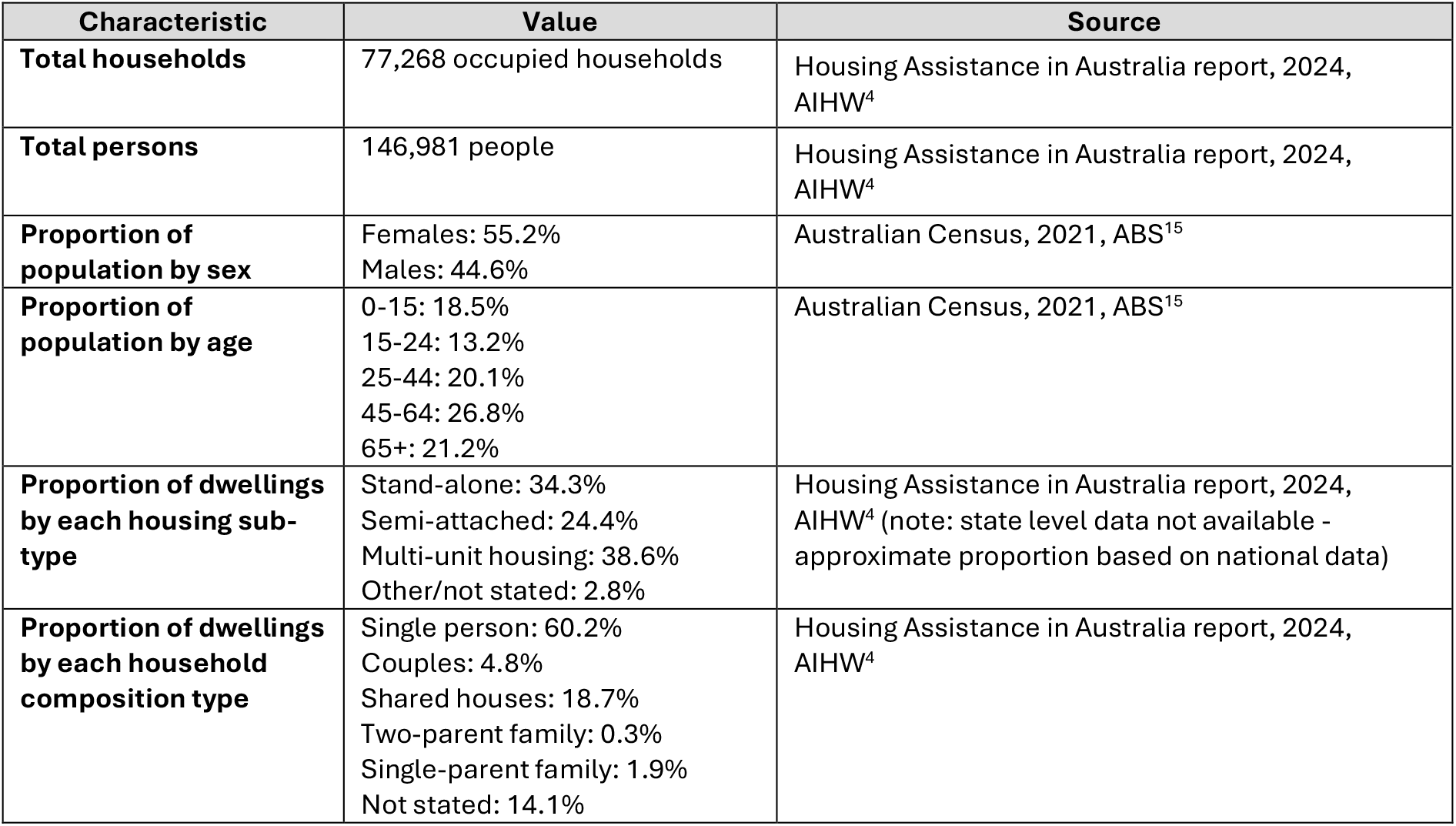
Characteristics of the social housing population in Victoria, Australia (2021-2023)

| Characteristic | Value | Source |
| --- | --- | --- |
| **Total households** | 77,268 occupied households | Housing Assistance in Australia report, 2024, AIHW<sup>4</sup> |
| **Total persons** | 146,981 people | Housing Assistance in Australia report, 2024, AIHW<sup>4</sup> |
| **Proportion of population by sex** | Females: 55.2%<br>Males: 44.6% | Australian Census, 2021, ABS<sup>15</sup> |
| **Proportion of population by age** | 0-15: 18.5%<br>15-24: 13.2%<br>25-44: 20.1%<br>45-64: 26.8%<br>65+: 21.2% | Australian Census, 2021, ABS<sup>15</sup> |
| **Proportion of dwellings by each housing sub-type** | Stand-alone: 34.3%<br>Semi-attached: 24.4%<br>Multi-unit housing: 38.6%<br>Other/not stated: 2.8% | Housing Assistance in Australia report, 2024, AIHW<sup>4</sup> (note: state level data not available - approximate proportion based on national data) |
| **Proportion of dwellings by each household composition type** | Single person: 60.2%<br>Couples: 4.8%<br>Shared houses: 18.7%<br>Two-parent family: 0.3%<br>Single-parent family: 1.9%<br>Not stated: 14.1% | Housing Assistance in Australia report, 2024, AIHW<sup>4</sup> |

#### We modelled three scenarios

1. Business-as-usual (BAU) – parameterised using projections of smoking prevalence and tobacco-disease rates, assuming continuation of current policies without further interventions.
2. Second-hand smoke eradication – smoking inside homes or multi-unit housing buildings ceases in 2025, with no change in smoking prevalence or intensity.
3. Complete smoking eradication – all people who smoke in the social housing population quit in 2025, with no further uptake.

*SHINE Tobacco* combines a Markov model of smoking behaviour with a proportional multistate lifetable (PMSLT)^12^ linking smoking trends to health outcomes across 31 smoking-related diseases. For this this study we expanded the platform with a Monte Carlo matrix model to simulate second-hand smoke exposure in multi-unit housing. The full model structure is shown in Supplementary Figure 1.

Model inputs included demographic data (age [0-110 years], sex [male, female], household structure [stand-alone, multi-unit housing]), housing characteristics (e.g., dwelling type, number of floors), smoking behaviours (uptake and quit rates, and smoking intensity), and disease-specific parameters (incidence, case fatality, and remission rates). Where direct data for the Victorian social housing population were unavailable, we used national estimates for the social housing population or for the most disadvantaged areas in Australia, identified using the Australian Bureau of Statistics (ABS) Socio-Economic Indexes for Areas (SEIFA).^16^ Smoking prevalence and behavioural trends were derived from the Household, Income and Labour Dynamics in Australia (HILDA) survey^17^ and the National Health Survey (NHS)^18^. Further detail on all inputs is provided in Table 2.

**Table 2.**
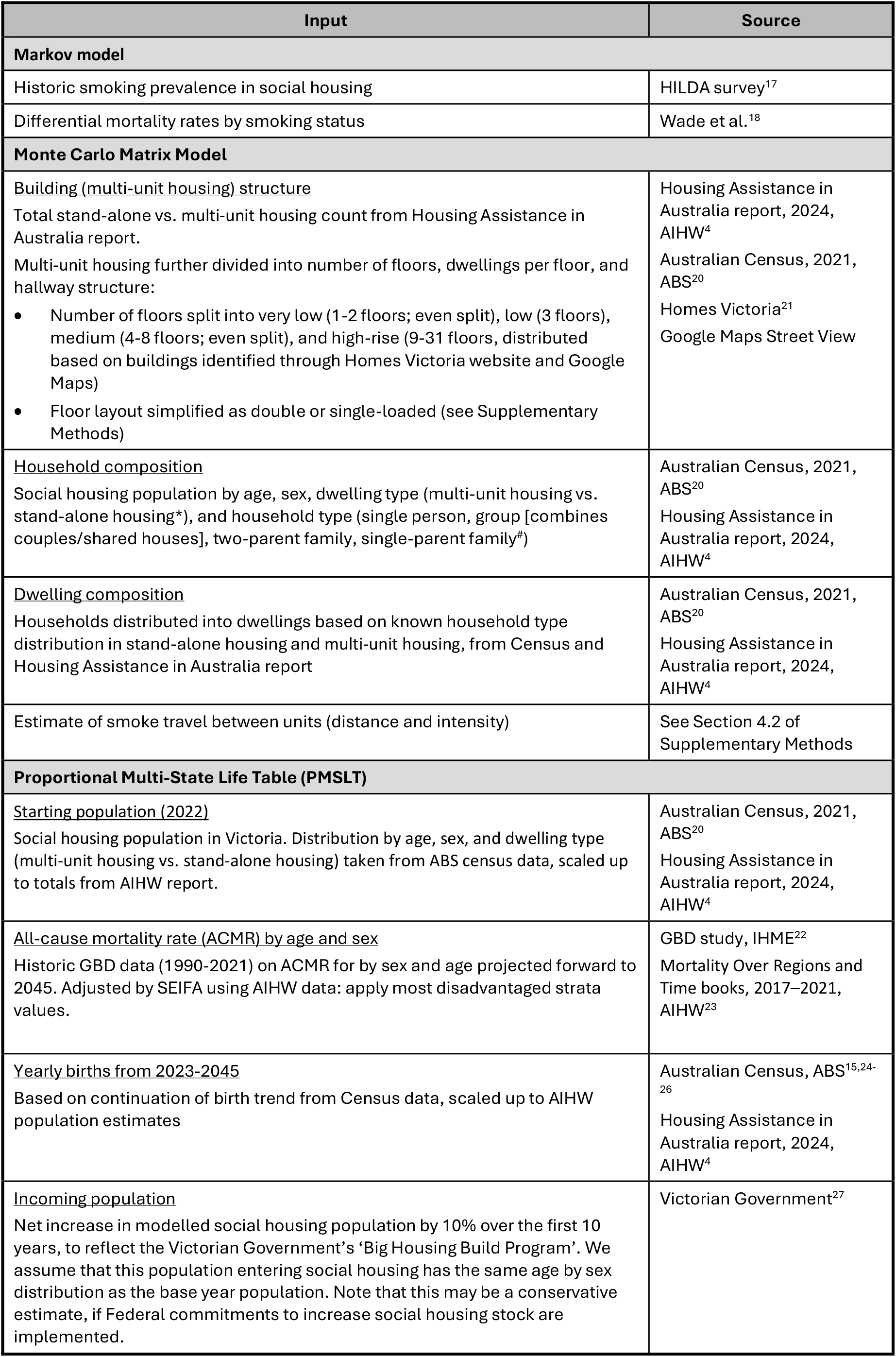
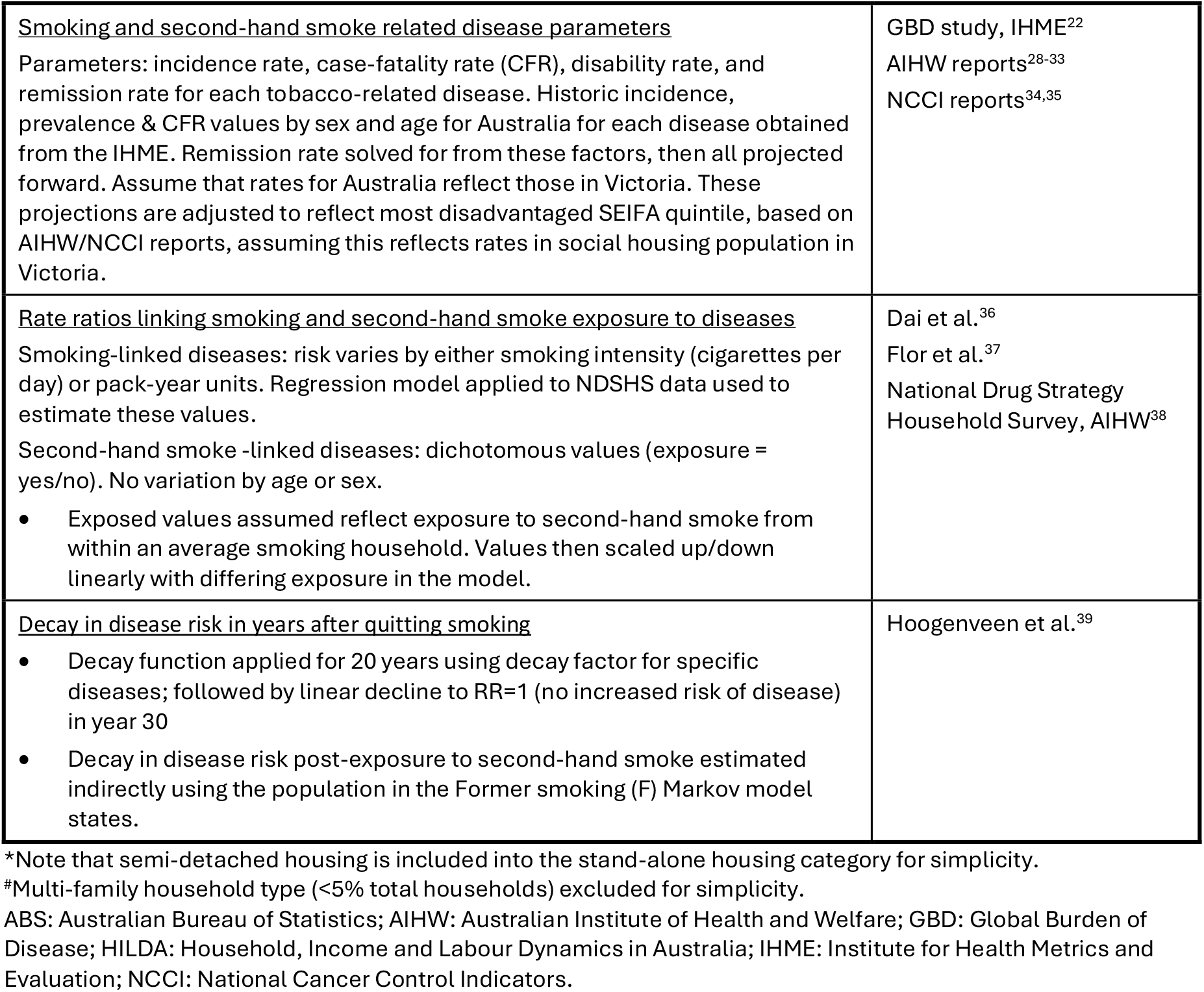
Model inputs.

### Smoking trends

Under the BAU scenario, smoking rates were estimated using a Markov model, with individuals categorised by sex and birth year into three states: Never smoked (***N***), Smokes daily (***S***), and Formerly Smoked daily (***F***) (Supplementary Figure 1). HILDA and NHS data indicate that smoking rates among social housing residents have remained stable in recent years (Supplementary Figure 2). We therefore assumed constant smoking uptake and cessation rates, using the most recent data for calibration (Supplementary Methods, Section 3.2).

### Second-hand smoke exposure

We developed a Monte Carlo matrix model to quantify second-hand smoke exposure from both within household smoke exposure and smoke drift in multi-unit housing. This model generates an infiltration matrix ***N***, representing the weighted proximity between each individual and potential sources of second-hand smoke.

#### The construction of *N* involves three components

- Dwellings in buildings – where a ‘building’ can be a single stand-alone dwelling or a multi-unit housing building. Each dwelling in multi-unit housing was assigned a three-dimensional coordinate: corridor position (***x***), side of the corridor (***y***), and floor level (***z***).
- Households – randomly assigned to dwellings based on composition (two-parent family, single-parent family, group, single person) and age-sex structure.
- Drift function – second-hand smoke exposure for each household was calculated based on dwelling proximity to smoking households. Maximum exposure occurred in smoking households (where 1 or more residents smoke). For other, second-hand smoke exposure declined with increasing horizontal and vertical distance from a smoking unit. Drift parameters were based on a rapid literature review and calibrated so that a non-smoking dwelling directly adjacent to a smoking dwelling received ∼31% less second-hand smoke than the smoking unit itself.^40^

The infiltration matrix was produced by applying the drift function to randomly generated buildings, averaging results over 10,000 iterations. Full parameterisation and sensitivity scenarios are described in the Supplementary Methods (Section 4.2 - 4.3).

### Combining health impacts

To estimate the health burden attributable to both direct smoking and second-hand smoke exposure, we linked the infiltration matrix ***N*** to the PMSLT. The PMSLT model^12^ includes an all-cause lifetable, and 31 disease-specific lifetables, allowing estimation of morbidity and mortality over time. Smoking prevalence and second-hand smoke exposure estimates were translated into changes in disease incidence using potential impact fractions (PIFs). The resulting changes in health outcomes were expressed in terms of health-adjusted life years (HALYs) gained^12^ and deaths averted.^41^

For active smoking, relative risks (RRs) were based on dose-response relationships in pack-years or daily cigarette equivalents, with risk for former smokers declining over 30 years post-cessation. For second-hand smoke, exposure levels were calculated by multiplying ***N*** by age- and sex-specific prevalence of current and former smoking, assuming that those who currently live with or near people who formerly smoked lived in the same setting during the period when those individuals smoked. Binary RRs from the Global Burden of Disease Study (GBD) were mapped to a continuous exposure scale anchored to the average number of smokers per smoking household in Australia. The same decay function was applied for former second-hand smoke exposure. Full methodological details are provided in the Supplementary Methods – Sections 4 and 5.

### Intervention scenarios

Two hypothetical tobacco control scenarios, both starting in 2025, were simulated and compared to BAU:

1. Second-hand smoke eradication – all household and multi-unit housing smoking ceases, with no change in smoking prevalence or uptake.
2. Complete smoking eradication – all daily smoking ceases, eliminating both direct smoking and second-hand smoke exposure.

These scenarios represent theoretical minimum risk exposure levels (TMREL)^42^ for second-hand smoke in the home, and for tobacco smoking overall.

The PMSLT was run through a Monte Carlo simulation of 2000 iterations, drawing uncertain parameters from their respective distributions. The model was run as an open cohort for 20 years, including births, as well as an increase in overall population living in social housing over the first 10 years reflecting recent government commitments to increase supply.^27^ Model outcomes (HALYs and deaths) under each scenario were compared to BAU (i.e., static smoking transitions and the consequent second-hand smoke exposure level). The results are presented as medians with a 95% uncertainty interval (UI). Age-standardised HALYs were calculated by direct standardisation to the 2001 Australian population.^43^

### Sensitivity analyses

We conducted a deterministic one-way sensitivity analysis, varying uncertain parameters to their 2·5^th^ and 97·5^th^ percentiles. We also conducted an additional analysis to explore alternative second-hand smoke drift assumptions, of:

- High drift – 19% reduction in exposure for directly adjacent non-smoking units.
- Low drift – 45% reduction for directly adjacent non-smoking units.
- No drift – second-hand smoke exposure only within smoking households.

Further details on parameterisation and these analyses are available in the Supplementary Methods (Section 4.2).

## Results

### Second-hand smoke eradication scenario

Compared to BAU, eradicating second-hand smoke exposure in the social housing population in Victoria was projected to avert approximately 600 (95% UI 500-700) deaths over 20 years (2025-2045) and lead to around 5,400 HALYs (95% UI 4,800-6,100) gained, equivalent to 86 HALYs per 100,000 person-years (95% UI 75-100). These health gains varied across sub-populations (Figure 1). Females gained 63% of the total HALYs, despite having slightly lower smoking rates than males (31% vs. 35%).

**Figure 1.**
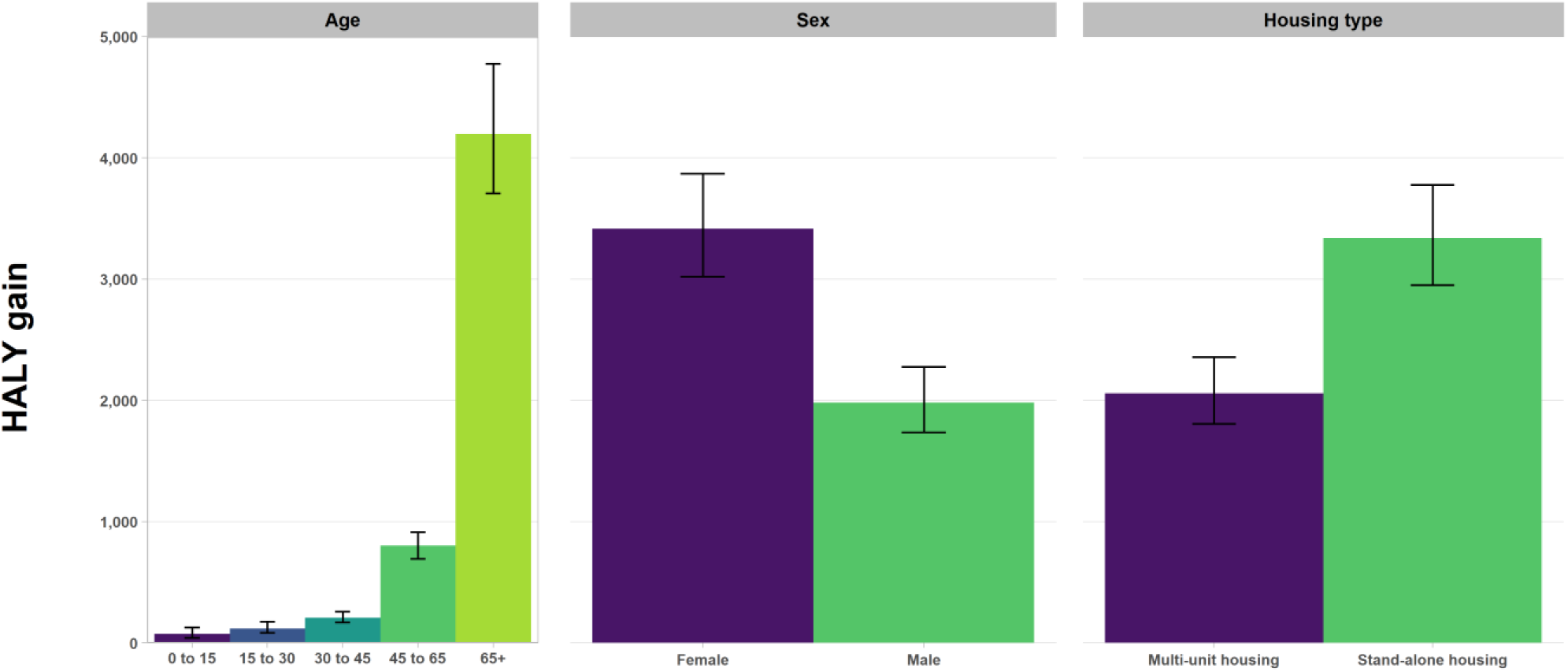
Health gained, in health-adjusted life years (HALYs), over 20-years under hypothetical second-hand smoke elimination scenario, by sub-group. Values shown as median HALY gain relative to business-as-usual, with 95% uncertainty interval as error bars.

The multi-unit housing sub-population received 38·1% of the total health benefit of second-hand smoke eradication, despite comprising only about 30% of the social housing population. This reflects the additional second-hand smoke burden from smoke drift. Within multi-unit housing, 54·4% of HALYs gained were due to the removal of smoke drift between units, with the remainder from eliminating exposure within smoking household (Supplementary Table 6-7).

### Smoking eradication scenario

In comparison to BAU, where smoking prevalence in the social housing population persists at 30-35% over the next 20 years, eradication all smoking was projected to result in 19,800 HALYs (95% UI 16,500-23,700) gained (or 430 per 100,000 person-years), and 1,430 premature deaths (95% UI 1,220-1,670) averted, over 20 years. Of the total health gain in this scenario, 27.3% of HALYs and 42.2% of deaths averted were attributable to the removal of second-hand smoke exposure in the home that would occur if all smoking ceased (Table 3). This proportion was greater in multi-unit housing (29·8% of HALYs, 44·4% of deaths averted) than in stand-alone housing (26·1% and 40·9%, respectively).

**Table 3.** Health gain under total second-hand smoke only vs. complete tobacco eradication, by sub-population, each relative to BAU, over 20-years (2025-2045).

| Outcome comparison | Total population | Multi-unit housing | Stand-alone housing |
| --- | --- | --- | --- |
| **HALY gain** |  |  |  |
| **Second-hand smoke eradication** | 5,400 (4,770 to 6,130) | 2,060 (1,810 to 2,360) | 3,340 (2,950 to 3,780) |
| **Total smoking eradication<sup>1</sup>** | 19,800 (16,500 to 23,700) | 6,920 (5,860 to 8,230) | 12,800 (10,600 to 15,500) |
| **% due to second-hand smoke removal** | 27.3% | 29.8% | 26.1% |
| **Deaths averted** |  |  |  |
| **Second-hand smoke eradication** | 603 (517 to 709) | 239 (204 to 282) | 365 (313 to 429) |
| **Total smoking eradication<sup>1</sup>** | 1,430 (1,220 to 1,670) | 538 (457 to 628) | 892 (757 to 1,040) |
| **% due to second-hand smoke removal** | 42.2% | 44.4% | 40.9% |

<sup>1</sup>Total smoking eradication results in eradication of second-hand smoke exposure.
BAU: business-as-usual; HALY: health-adjusted life year.

### Sensitivity analysis

The tornado plot in Supplementary Figure 6 shows the contribution of model inputs to uncertainty in HALYs gained. No single input dominated total uncertainty in HALYs gained. The largest contributors were lung cancer incidence rates in BAU (19·3% of total uncertainty), followed by chronic obstructive pulmonary disease (COPD) incidence rates in BAU (18·7%), and ischaemic heart disease (IHD) incidence rates (13·9%).

The sensitivity analysis showed that health gain estimated from tobacco eradication was moderately sensitive to smoke drift assumptions (Supplementary Tables 6-7). Altering the rate of decay of smoke drift between households (multi-unit housing only) resulted in a change in deaths averted in the multi-unit housing sub-population under second-hand smoke eradication by up to 8·4% compared to the main analysis (Supplementary Table 7).

## Discussion

This study provides new evidence on the potential health gains from reducing both direct smoking and second-hand smoke in social housing, with a particular focus on the contribution of smoke drift in multi-unit housing. Our model estimated that eliminating second-hand smoke exposure in Victoria’s social housing population could result in 5,400 (95% UI 4,800-6,100) HALYs gained, and 600 (95% UI 500-700) deaths averted over a 20-year period. The model also shows that second-hand smoke elimination would account for approximately 27-42% of the total potential benefit of complete smoking eradication. Under this scenario – where all residents who smoke quit in 2025 and no further uptake occurred – an estimated 19,800 (95% UI 16,500-23,700) HALYs could be gained, and 1,400 (95% UI 1,200-1,700) deaths averted over 20 years.

Within the social housing population, there were marked differences in health gain. Females gained a larger share of health benefits from second-hand smoke elimination than males, despite slightly lower smoking prevalence (31% vs. 35% in the base year). This is consistent with their higher overall representation in social housing (57% of social housing residents in Victoria are females) and with additional second-hand smoke -related health impacts, such as breast cancer.^44^ The disproportionate health gains estimated for multi-unit housing residents reflect both their higher baseline exposure to second-hand smoke and the unique contribution of smoke drift, with over half of health gains (54·4% HALYs gained, and 61·4% deaths averted) being attributable to removing smoke drift between units rather than exposure from smoking within the household. These findings highlight the importance of smoke drift, which is often overlooked in burden of disease studies; these studies typically estimate second-hand smoke exposure from self-reported measures of living with a person who smokes and do not account for inter-unit transfer in apartment buildings.

Our results are consistent with previous evidence showing a high burden of tobacco-related harms among social housing residents and are consistent with the higher smoking rates experienced by this population in Australia^45^ and globally^46^. Our results also highlight the stark health inequality due to tobacco when compared with the broader Australian population.

The modelled scenarios reflect hypothetical “maximum impact” cases. The second-hand smoke elimination scenario assumed that all indoor smoking ceased and full compliance was maintained. While no real-world policy is likely to achieve such immediate and complete adherence, the scenario provides an estimate of the upper bound of potential benefits. Further, the smoking eradication scenario demonstrates the full scale of benefit possible from maximally effective tobacco control in this setting. Achieving either complete eradication of tobacco use or parity in smoking prevalence between social housing residents and the broader population would require substantial changes in policy and resources. Internationally, the most relevant example is the US Department of Housing and Urban Development’s 2018 federal rule banning smoking in all public housing buildings and within a set distance from entrances.^8^Implementation and enforcement of the rule, including provision of cessation support for those who smoke in parallel to the ban, has been varied.^9^ In the Australian context, the likelihood of adopting a smoke-free housing policy or similar at the national level in the near-term is unclear.

Our findings suggest that targeted second-hand smoke reduction strategies could achieve meaningful health gains even without large reductions in smoking prevalence. Potential approaches identified in the literature include implementing smoke-free housing policies that cover private dwellings in multi-unit housing^47^, integrating smoking cessation programs into tenancy and housing services to improve smoke-free policy effectiveness, and establishing monitoring systems to measure second-hand smoke exposure and evaluate the effectiveness of interventions^48^. These measures, however, must be designed with careful attention to potential unintended consequences, such as eviction risk, compliance challenges, and stigma. In a qualitative study in affordable housing, more than half of smoke-free policy violations resulted in lease terminations or evictions.^49^ These issues are particularly salient in populations experiencing housing insecurity and other forms of structural disadvantage, reinforcing the need for supportive, non-punitive approaches. There are also ethical considerations that come with regulating smoking behaviour in private living spaces, especially for populations with housing needs.^50^

To our knowledge, this is the first study to link a spatial model – incorporating the physical layout of dwellings and the relative location of smoking households – to a PMSLT model to estimate long-term health impacts of changes in smoking and second-hand smoke exposure. The Monte Carlo matrix quantified second-hand smoke exposure as a continuous variable considering both the number of nearby smoking households and their horizontal and vertical proximity. This approach addresses the limitations of self-reported second-hand smoke exposure, which generally underestimate true exposure levels.^51,52^ Our modelling framework can also be adapted for other high-density living contexts where residents are exposed to airborne hazards (including tobacco smoke, traffic-related or industrial emissions) from multiple nearby sources. With apartment living becoming increasingly common in Australia^5^, this type of modelling could help quantify the potential benefits of interventions beyond smoke-free housing policies.

Our study has several limitations. First, we assumed that the health status of the Victorian social housing population was equivalent to that of the most disadvantaged SEIFA quintile in Australia. while this was a pragmatic approach, it may not fully capture the health and demographic profile of the social housing population.

Second, when calibrating the Markov model, we assumed that increases in smoking prevalence with age were entirely due to uptake and decreases were entirely due to cessation (or smoking-related death). This assumption could be problematic if there is differential movement into or out of social housing by smoking status. Given the known association between smoking and other factors linked to social housing need – such as income level or mental illness – those who do not smoke may be more likely to leave the social housing population than those who do smoke, or conversely, people who smoke may be more likely to enter. These patterns could produce an artificial increase in apparent smoking uptake or a decrease in cessation rates in the calibration process. Longitudinal studies that incorporate movement into and out of social housing, priority group allocation, and additional socio-demographic variables, such as ethnicity, mental health status, and substance use, would allow more nuanced projections of this population.

Third, the incoming population (modelled to reflect the anticipated growth in social housing stock in Victoria^27^) was assumed to have the same characteristics – including sex-by-age distribution and smoking prevalence – as the baseline population. This may not be reasonable, given that there are priority groups for social housing allocation that may not reflect historical allocation policies underlying the current population profile.

Fourth, our model lacks heterogeneity beyond sex, age, and housing sub-type. Largely as a result of the ongoing impacts of colonisation, the First Nations population is disproportionally represented in social housing in Australia^4^ and has higher rates of both tobacco smoking, and second-hand smoke exposure, the latter often related to overcrowding in housing)^53,54^. Other chronic factors – including substance use and mental health disorders – are also over-represented in social housing populations and are associated with a higher prevalence of tobacco use than the population average.^55^ Future modelling, and policy development, should include such factors explicitly.

Finally, there was a lack of quantitative data on smoke infiltration between units in multi-unit housing. We therefore use a simplified approach to estimate smoke drift, which did not differentiate between potential transmission routes (e.g., balconies, open windows, hallways) or account for seasonal variations in smoke drift. The uncertainty in this input parameter was addressed by scenario analyses that altered the decay rate in the smoke drift function (Supplementary methods, Section 4.2). Increasing or decreasing the strength of smoke drift altered the modelled health outcomes by 5·3-8·4% (Supplementary Tables 6-7). When smoke drift was completely excluded from estimates, the health gain from removing second-hand smoke in multi-unit housing more than halved. Future research is needed to provide empirical data on smoke drift under different building designs, occupancy patters, and climatic conditions to inform more accurate modelling.

## Conclusion

This modelling study of second-hand smoke exposure in social housing – a setting characterised by both high smoking prevalence and second-hand smoke -density living – identified two key findings. First, eliminating second-hand smoke exposure could account for a substantial proportion – between 27 and 42% – of the total health gain achievable through complete elimination of tobacco use and exposure. Second, in multi-unit housing, over 50% of potential health gain from removing second-hand smoke exposure may come from elimination of smoke drift between dwellings specifically. Together, these results highlight the importance of addressing second-hand smoke as a distinct source of preventable disease burden in socioeconomically disadvantaged populations. Our modelling provides a strong empirical rational for policies to effectively address second-hand smoke exposure in social housing. Future research should focus on improving empirical estimates of smoke drift, incorporating more detailed socio-demographic and health profiles.

## Data Availability

Input data used in this study were taken from publicly available sources (exception: Unit-level Household, Income and Labour Dynamics in Australia (HILDA) survey data was obtained via a request from the Australia Data Archive). Model code available upon reasonable request to the authors.

## Acknowledgements

Funding was received through the NHMRC Centre of Research Excellence on Achieving the Tobacco Endgame (CREATE; GNT1198301).

This research was supported by The University of Melbourne’s Research Computing Services and the Petascale Campus Initiative.

## Notes

### Author Declarations

Unit-level Household, Income and Labour Dynamics in Australia (HILDA) survey data was obtained via a request to the Australia Data Archive (ethics approval not required for ADA access).

