## Supplementary Material for "Estimating the harms from smoking and second-hand smoke exposure in social housing: a modelling study"

#### Contents

**Supplementary Methods** ... 2

1. Model overview ... 2
2. Study population ... 2
3. Markov model ... 2
   3.1 Markov model overview ... 2
   3.2 Input data ... 3
   3.3 Model calibration ... 4
4. Monte Carlo matrix model ... 6
   4.1 Matrix model overview ... 6
   4.2 Data inputs ... 7
   4.2 Second-hand smoke infiltration ... 7
   4.3 Modelling second-hand smoke intensity as a function of smoking rates ... 9
5. PMSLT ... 10
   5.1 PMSLT overview ... 10
   5.2 PMSLT data sources ... 10
   5.3 Potential Impact Fraction ... 11
   5.4 Smoking-disease associations ... 12
   5.5 Second-hand smoke-disease associations ... 14

**Supplementary Results** ... 15

**References** ... 17

#### Supplementary Methods

#### 1. Model overview

This technical appendix details the construction of a three-component simulation model system used to estimate the health impacts of direct-smoking and second-hand smoke exposure in a high-density, at-risk population. The model builds on an existing tobacco simulation platform, *SHINE Tobacco*, consisting of a Markov model and proportional multi-state lifetable (PMSLT). An earlier iteration of *SHINE Tobacco* has been used in previous research simulating 'tobacco endgame' interventions at a country-level in Aotearoa/New Zealand. (1-4) The PMSLT has also been applied more broadly to other risk factor/disease models. (5-7)

For this analysis, the platform was expanded to incorporate a pilot model of second-hand smoke exposure within an individual's home. We developed a novel approach that estimates second-hand smoke exposure as a continuous variable, varying according to the number of sources of exposure and the distance from each source. Importantly, this component is used to quantify second-hand smoke drift between dwellings in multi-unit housing.

The purpose of each model component is summarised as follows:

1. Markov model
   - Purpose: generate smoking uptake and quit rates and resulting smoking prevalence in the model population, under 'business-as-usual' (BAU) and intervention scenarios
2. Monte Carlo matrix model
   - Purpose: generate the model population and define the amount of possible second-hand smoke exposure to and from different dwellings in multi-unit housing, under BAU
3. Proportional Multi-State Lifetable (PMSLT)
   - Purpose: estimate health impacts from 31 smoking-related diseases and eight second-hand smoke-related diseases under intervention scenarios, in comparison to BAU, based on smoking rates from the Markov model, and contact between individuals from the matrix model

#### 2. Study population

The model is piloted using the social housing population of Victoria, Australia. This population consists of around 83,000 dwellings and 147,000 people as of 2023 (approximately 20% of the country's total social housing population). (8) Social housing in Australia includes both public housing provides by state governments, community housing run by non-government organisations, state owned and managed Indigenous housing (not operational in Victoria), and Indigenous community housing. (8) These housing options are available for people with a low income, with priority access given to specific groups, for example those who are currently homeless or those people a disability.

Approximately 39% dwellings in social housing are flats/apartments (as of 2023 (8)), higher than the average of 16% across all private dwellings. (9) Melbourne has the highest number of social housing dwellings of any region in Australia. This includes over 40 high-rise apartment buildings, currently undergoing redevelopment as part of a 10-year plan to improve and expand social housing in the state of Victoria. (10,11)

Daily smoking rates in the social housing population are significantly higher than the population average (34% (12) compared to 10% (13) for 15+ year olds) and smoking within the home is more common in this setting. There is therefore an increased risk of both direct-smoke and second-hand smoke related health harms for people living in social housing. Currently, national-level laws do not exist in Australia that prohibit smoking in private residences within apartment buildings. This creates a potentially greater risk for the sub-population living in apartments (i.e., multi-unit housing) of smoke infiltration from neighbouring units in this setting.

#### 3. Markov model

##### 3.1 Markov model overview

The structure Markov model is presented in Supplementary Figure 1. The three main states represent smoking-related behaviours: Never smoked (***N***), currently Smokes daily (***S***), and Formerly smoked daily (***F***). Birth cohorts move between these three states in yearly cycles. An additional Death state ($\boldsymbol{D}$) is included to incorporate differential all-cause mortality by smoking status. The F state is split into 30 states in a 'tunnel', where movement from state $\boldsymbol{F_n}$ to $\boldsymbol{F_{n+1}}$ or $\boldsymbol{D}$ occurs with 100% probability until $\boldsymbol{F_{30}}$ is reached (the final $\boldsymbol{F}$ state from which there can be no further movement except to state $\boldsymbol{D}$). This structure allows for a gradual decrease in disease risk following smoking cessation (see section 5.3). All quit events are final – there is no relapse.

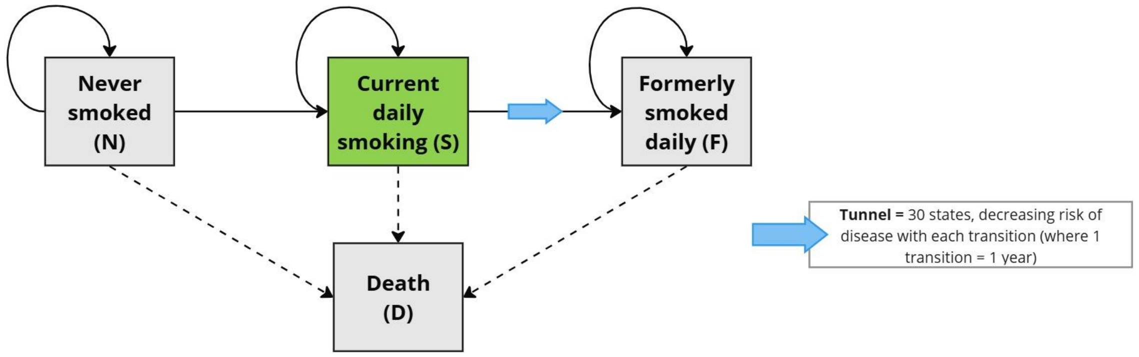


**Supplementary Figure 1. Smoking Markov model.**

##### 3.2 Input data

The inputs for the Markov model are summarised in Supplementary Table 1.

**Supplementary Table 1. Markov model data inputs**

| Input | Source | Detail |
|---|---|---|
| Historic smoking prevalence in social housing | Household, Income and Labour Dynamics in Australia (HILDA) survey (14) [1] | Sex by age-grouped data in 2021-2022 converted to single year of age values using cubic spline interpolation. |
| Differential mortality rates by smoking status | Wade et al. (15) | Differential mortality by smoking status (current, former, never) by age, taken from an analysis of the Australian 45 and up study data. |
| Starting population (2022) | Australian Census, 2021, ABS (16)<br>Housing Assistance in Australia report, 2024, AIHW (8) | Population disaggregated by age, sex, and dwelling sub-type (multi-unit housing vs. separate dwellings) |
| Birth projections | Australian Census, ABS (16-19)<br>Housing Assistance in Australia report, 2024, AIHW (8) | Historic Census data scaled up to AIHW population estimates. Linear trend applied for 10 years, then birth count held constant for subsequent years. |
| Migration projections | Victorian Government (11) | Assume no net international migration into/out of social housing population.<br>Apply net influx of people into social housing from the base year, over a 10-year period, to increase the total social housing population by 10% from the base year value. This is based on the Victorian Government's 'Big Housing Build Program', which specifies that social housing stock will be increased by 10%. We assume that the population entering social housing has the same age by sex distribution as the base year population. |
| All-cause mortality rate projections | GBD study, IHME (20)<br>Mortality Over Regions and Time books, 2017–2021, AIHW (21) | Historic GBD data on ACMR for by sex and age projected forward.<br>Adjusted by SEIFA using AIHW data: apply most disadvantaged strata values. |

[1]HILDA data obtained via request from the Australia Data Archive.
ABS: Australian Bureau of Statistics; AIHW: Australian Institute of Health and Welfare; GBD: Global Burden of Disease; IHME: Institute for Health Metrics and Evaluation

The population for the Markov model (and the PMSLT, discussed in the next section) is defined by yearly births, migration, and mortality rates by sex, age, and housing sub-type. The Markov model is calibrated with 2022 as the base year, given that this is the last year of available smoking prevalence data (Supplementary Table 1).

Historic data on daily smoking prevalence in the Australian social housing population are limited. Two datasets collect information on both smoking and housing – the Household, Income and Labour Dynamics in Australia (HILDA) survey (22), and National Health Survey (NHS). (13) Daily smoking prevalence in the social housing population from these two sources is presented in Supplementary Figure 2. Linear trend lines through datasets show a minor positive linear trend with time.

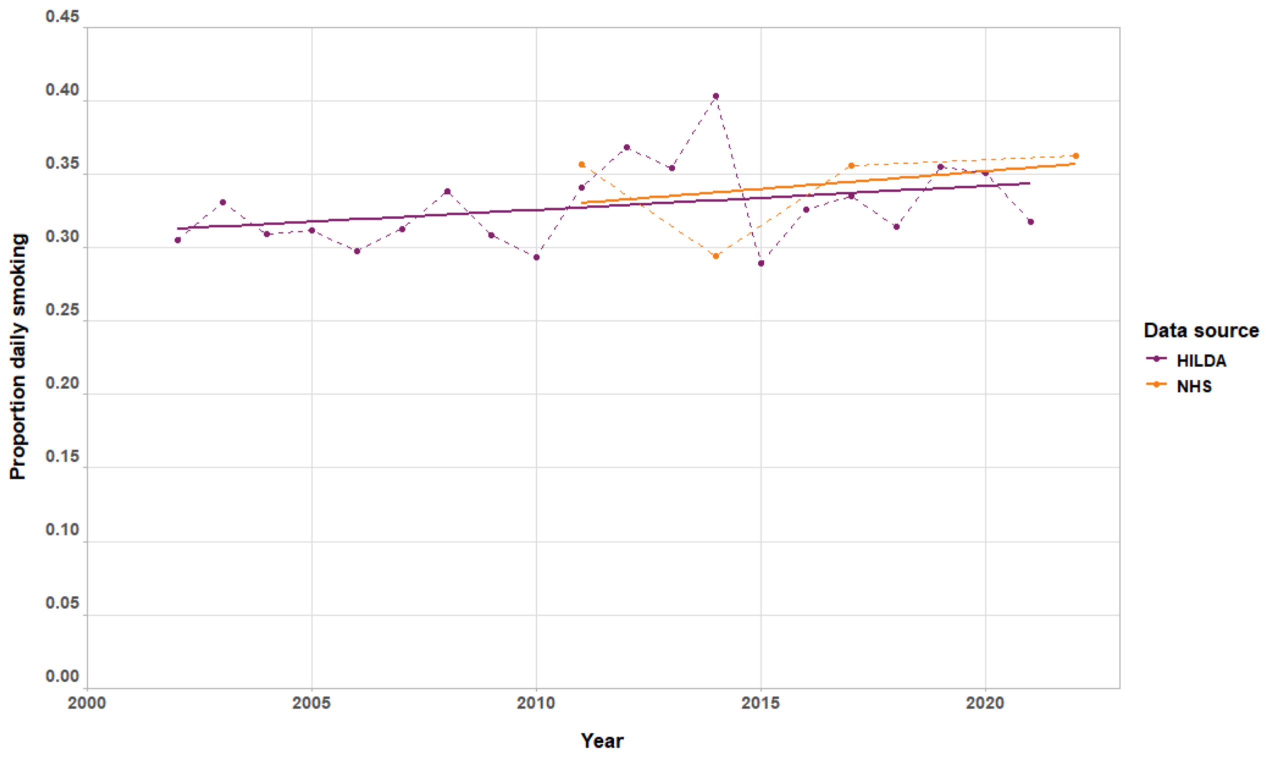


**Supplementary Figure 2. Historic daily smoking rates in social housing, Australia.**
Solid lines represent linear trend lines by year for each source. HILDA data sourced from the Australian Data Archive (by request) (14); NHS data obtained from Australian Bureau of Statistics TableBuilder Platform. (12, 23-25) HILDA: Household, Income and Labour Dynamics in Australia; NHS: National Health Survey.

##### 3.3 Model calibration

We used HILDA data, obtained as unit-level data by request from the Australian Data Archive (ADA). (14) The HILDA survey is a yearly panel study of Australian households, initiated in 2001. (23) The study has collecting smoking data since wave 2. Daily smoking prevalence by sex and age group (15-24, 25-40, 40-55, and 55+ years) was derived from the cross-sectional sample, specifically for the social housing sub-group, using the *Survey* package (26) in R to account for the complex survey design. Due to the small sample sizes, it was not possible to disaggregate yearly prevalence estimates into finer age groups, which would have allowed more accurate estimation of birth cohort trends (i.e., smoking uptake and cessation as a cohort ages).

The calibration process was as follows:

1. **Interpolation and cohort conversion**
   Age grouped data from the most recent two waves (2021-2022) of the HILDA survey were converted into single years of age using cubic spline interpolation, then converted to values by birth cohort.
2. **Initial transition rate estimation**
   The differences in smoking prevalence from 2021-2022 for each synthetic birth cohort were converted to an initial estimate of uptake and cessation transition rates. A positive change was treated as uptake and a negative change as cessation. E.g., if the smoking prevalence in 2021 for male 20-year-olds was 10%, and in 2022 for male 21-year-olds (i.e., for the same birth cohort), was 12%, then the uptake rate is $(0.12 - 0.10)/0.10 = 20\%$.
3. **Optimisation**
   Uptake and cessation transition rates calibrated to smoking prevalence targets using a mathematical optimisation model. The model uses a simulated annealing algorithm (27) to explore a parameter space of all possible combinations of uptake and cessation transitions by sex, age, and year, aiming to reduce the difference between the modelled output and the target prevalence. The target was the 2022 smoking prevalence by age and sex. Given the lack of change in smoking prevalence over time in social housing in Australia (Supplementary Figure 2), the prevalence values for 2022 were also applied to each future year of the model (2023-2044), assuming stable prevalence. The algorithm starts with the initial transitions from step 2, and iteratively explores alternative values, minimising the difference between the target prevalence values, and the prevalence values produced from the modelled uptake and quit transitions. Smoking uptake rates ($\boldsymbol{N} \rightarrow \boldsymbol{S}$ transition) were constrained so that uptake can only occur from ages 15-40, and cessation rates ($\boldsymbol{S} \rightarrow \boldsymbol{F}$ transition) constrained to occur only for ages 35+. Transitions to state D were fixed (Supplementary Table 1) to ensure that decreases in smoking prevalence as a cohort age account for smoking-attributable deaths as well as cessation.

Supplementary Figure 3-4 show the projected annual daily smoking rates resulting from the calibrated transitions. It should be noted that while the calibration target specified stable smoking rates into the future (at the 2022 level by sex and age), smoking rates at the population-level show a slight variation over time due to changes in the population structure in future years.

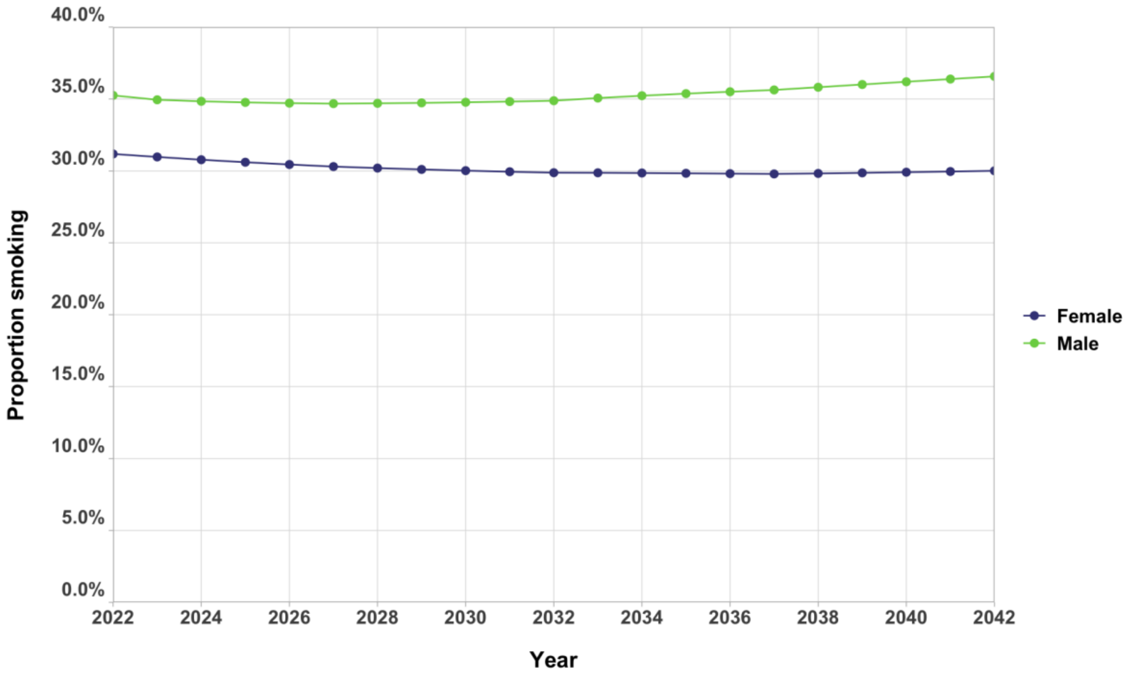


**Supplementary Figure 3. Calibrated daily smoking prevalence by sex.**

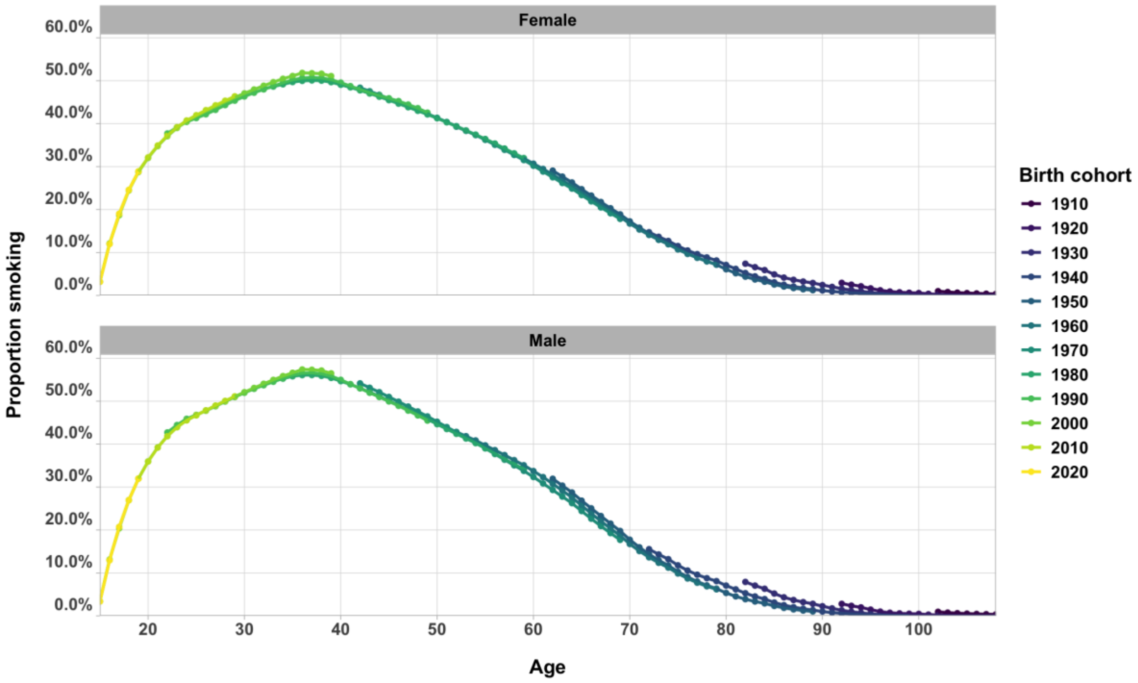


**Supplementary Figure 4. Calibrated daily smoking prevalence by birth cohort.**
Smoking uptake peaks around age 35-40 for all birth cohorts, with the downward slope beyond this age representing the combination of quitting smoking and smoking-related mortality. Birth cohorts are grouped into 5-year cohorts (e.g., 1910 = born from 1910-1914).

It was assumed that there is no differential movement in/out of social housing by smoking status, as data was not available to distinguish between changes in smoking prevalence due to migration vs. smoking cessation. This is an area for further research.

### 4. Monte Carlo matrix model

#### 4.1 Matrix model overview

In our model, we define second-hand smoke exposure as occurring within one's home and arising either from someone living in the same dwelling who smokes or from someone who smokes in a dwelling within the same building (within a defined proximity). Exposure is represented as a continuously, allowing:

a) aggregation of exposure from multiple sources (e.g., living with two people who smoke, or having multiple neighbouring dwellings that are smoking households); and
b) variation of exposure according to the proximity of other smoking dwellings to the individual receiving exposure.

The Monte Carlo matrix model simulates the distance between individuals and dwellings in the social housing population to estimate varying levels of second-hand smoke exposure on a continuous scale.

The matrix model is constructed with three components:

1. Households – defined by the number of people and sex by age compositions, residing in dwellings. Households were categorised into the main household compositions defined by the Australian Bureau of Statistics (ABS): two-parent families with variable child count, single-parent families with variable child count, single individuals, or groups (which includes both couples without children, and shared

households). Other composition types, such as multiple family households and unclassified households, contribute to <5% of total households and were excluded for simplicity. (28)

2. Dwellings in buildings – buildings range from a single dwelling (separate or stand-alone houses) to multi-unit housing buildings with up to 30-floors. Multi-unit housing building were further categorised number of floors, number of dwellings per floor, and dwelling layout: double loaded (central hallway with units on either side) or single-loaded (units on one side of the hallway only).
3. Exposure function – defines the level of second-hand smoke an individual could be exposed to, based on the characteristics of the dwelling and building an individual resides in.

The model includes second-hand smoke exposure from other dwelling within the same building. This means that residents in separate or stand-alone dwellings can be exposed only to second-hand smoke from other occupants in the same dwelling. The model does not account for variation in second-hand smoke exposure due to building features such as balconies, differing ventilation systems, or open doors/windows within multi-unit housing buildings.

##### 4.2 Data inputs

The inputs of the matrix model are shown in Supplementary Table 2.

**Supplementary Table 2. Monte Carlo matrix model data inputs**

| Input | Source | Detail |
|---|---|---|
| Multi-unit housing building count and structure (number of floors, dwellings per floor, floor layout) | Australian Census, ABS (9)<br>Google Maps<br>Homes Victoria (10) | • High rise dwelling count available directly from Homes Victoria<br>• Low and mid-rise building count estimated indirectly using census data on the number of dwellings, and assigning these dwellings to buildings of varying sizes<br>• Number of floors was determined through a combination of Homes Victoria documents (to get a sample of low and mid-rise buildings) and Google Maps Street View (to determine the number of floors for high rise buildings)<br>• Floor layout simplified as double or single-loaded |
| Social housing population by age, sex, and dwelling type (multi-unit housing vs. separate housing) | Australian Census, ABS (28)<br>Housing Assistance in Australia report, 2024, AIHW (8) | • AIHW data used for total count.<br>• Separated by age and sex using proportions from ABS census data. |
| Dwelling and household structure | Housing Assistance in Australia report, 2024, AIHW (8) | • Household type: single person, group (combining couples/share houses), single-parent family, couple family<br>• Dwelling types: multi-unit housing vs. stand-alone housing |
| Estimate of smoke travel between dwellings (distance and intensity) | Assumption (based on literature review) | See Section 4.2 |

ABS: Australian Bureau of Statistics; AIHW: Australian Institute of Health and Welfare

##### 4.2 Second-hand smoke infiltration

###### *4.2.1 Literature search*

A rapid literature review was conducted on 29/08/23, during model conceptualisation, to identify studies assessing second-hand smoke exposure levels (using quantitative measures such airborne nicotine levels, salivary/serum cotinine, or $PM_{2.5}$) in multi-unit housing. The search was initially limited to the social housing setting, but returned few results, so it was widened to all multi-unit/apartment settings. The search was updated on 30/03/25 to identify any additional studies published during model development.

The search strategy is summarised in Supplementary Table 3.

**Supplementary Table 3. Rapid literature search details.**

| Database | Search terms | Results [1] |
|---|---|---|
| PubMed | Pubmed: (("multi-unit hous*"[Title/Abstract]) OR ("multi unit hous*"[Title/Abstract]) OR ("multiunit hous*"[Title/Abstract]) OR (apartment[Title/Abstract])) AND (("secondhand smoke"[Title/Abstract]) OR ("second hand smoke"[Title/Abstract]) OR ("environmental tobacco"[Title/Abstract]) OR ("passive smok*"[Title/Abstract]) OR ("tobacco pollution"[Title/Abstract]) OR ("tobacco smoke pollution"[Title/Abstract])) AND (exposure OR level OR PM2.5 OR measure*) | 94 |
| Scopus | TITLE-ABS-KEY ( "multi?unit hous*" OR apartment ) AND TITLE-ABS-KEY ( "second?hand smoke" OR "environmental tobacco" OR "passive smok*" OR "tobacco pollution" OR "tobacco smoke pollution" ) AND ( exposure OR level OR pm2.5 OR measure* ) | 78 |
| After removing duplicates | | 143 |

[1]Including results from updated search on 30/03/25

Most identified studies were from the United States, followed by South Korea. One Australian study was found, (29) though it relied on self-reported second-hand smoke exposure. Field studies using $PM_{2.5}$ or airborne nicotine to estimate second-hand smoke in smoke-free units (dwellings with only people who do not smoke), generally did not stratify the study population by distance between smoking and non-smoking apartments. These studies suggested that nicotine exposure is greater on higher floors, with the difference more pronounced in winter. Gill et al. concluded that this is likely because nicotine moves in gaseous and particle phases of second-hand smoke and can rise with airflow. (30)

The few studies that have stratified by distance found that smoke moved into horizontally adjacent apartments, into hallways between units, as well as vertically adjacent apartments (moving up but not down). Levels of nicotine or $PM_{2.5}$ varied between studies, influenced by season (30, 31), proximity between units, building ventilation, and the amount smoked. (32) King et al. found that two out of 14 smoke-free units in a multi-unit housing complex had measurable second-hand smoke exposure (via $PM_{2.5}$) (32) and both were next to, or diagonally adjacent to smoking units. In the same study, $PM_{2.5}$ levels were more commonly detected, and at higher averages, in hallways than in non-smoking homes.

Ang et al. estimated $PM_{2.5}$ levels in Singapore apartments, comparing non-smoking households with smoking neighbours to smoking households. (33) $PM_{2.5}$ was reduced by 53%-73% in the former group compared with smoking households.

A single building simulation by Fabian et al. (34) modelled smoke travel in multi-unit housing incorporating dwelling size and building structure, temperature, and window door/opening. The authors found that 99% of the smoke infiltration from other apartments was coming from directly adjacent dwellings, both vertically and horizontally. (34)

While our literature search did not provide a direct quantitative risk estimate for second-hand smoke travel at varying distances, it informed several broad rules for out model:

- Smoke can travel from a given dwelling to adjacent dwellings (directly next to and diagonally) on the same floor, as well as directly to the dwelling above. Exposure in the adjacent dwellings is less than in the source dwelling.
- Some smoke remains in hallways on floors what at least one dwelling is occupied by a person who smokes indoors. A hallway exposure factor is therefore applied to all dwellings on the same floor of someone who smokes, as hallways are not explicitly modelled.

*4.2.2 Second-hand smoke infiltration function*

Informed by the literature search, we developed the following second-hand smoke infiltration function. We model each dwelling in a multi-unit housing block as having integer coordinates $(\boldsymbol{x}, \boldsymbol{y}, \boldsymbol{z})$, where $\boldsymbol{x}$ is distance along the hallway, $\boldsymbol{y}$ is the side of the hallway (either 0 or 1), and $z$ is the floor of the dwelling. For two dwellings A and B, let $(\boldsymbol{x}, \boldsymbol{y}, \boldsymbol{z})$ be the vector from A to B. Then the second-hand smoke that is received in B per person who smokes in A is defined by $\boldsymbol{f}\colon \mathbb{Z}^3 \rightarrow [0,1]$, with

$$f(x,y,z) = \begin{cases} (h^{-\max\{|x|,|y|\}} \times (1-w)) + w, & if\ z = 0 \\ v \times h^{-\max\{|x|,|y|\}}, & if\ z = 1 \\ 0, & if\ z \leq -1\ or\ z \geq 2 \end{cases}$$

where $h \geq 1$ is a horizontal decay factor, $v \in [0,1]$ a vertical decay factor, and $w \in [0,1]$ a hallway effect. So, all dwellings on the same floor receive at least $w$ second hand smoke along the hallway, and smoke infiltration drops by a factor of $h$ as dwellings becoming increasingly distant from a source of second-hand smoke.

The factor $\boldsymbol{h}$, $\boldsymbol{w}$, and $\boldsymbol{v}$ were set based on limited knowledge on second-hand smoke travel between dwellings in multi-unit housing, as determined in the rapid review (Section 4.2.1), with the central estimates shown in Supplementary Table 4. The decay estimates used for the main analysis were set at a level that resulted in a 31% reduction in second-hand smoke exposure from a smoking unit to a directly adjacent non-smoking unit, roughly based on the findings of Ang et al. (33) Lower and upper estimates were used in sensitivity analyses, to test how sensitive the model output was to specification of smoke drift. For the lower estimate, second-hand smoke exposure reduced from a smoking unit to an adjacent non-smoking unit by 50%, and under the upper estimate, by 20%.

**Supplementary Table 4. Dwelling-dwelling influence function parameters.**

| Variable | Central estimate | Lower estimate | Upper estimate |
|---|---|---|---|
| $h$ – horizontal decay factor | 1.5 | 2 | 1.25 |
| $v$ – vertical decay factor | 0.2 | 0.05 | 0.4 |
| $w$ – hallway effect [1] | 0.1 | 0.1 | 0.1 |

[1]Hallway effect not modified in sensitivity analyses, only decay factors.

##### 4.3 Modelling second-hand smoke intensity as a function of smoking rates

With $\boldsymbol{f}$ in hand, we can theoretically determine the second-hand smoke exposure experienced by individuals within multi-unit housing. This would be done as follows.

- Fill the multi-unit housing with households, generated as per Section 4.1.
- Randomly assign individuals a smoking status, with prevalence matching their sex and age as determined in Section 3.
- For each person who smokes, iterate over all individuals living in the multi-unit housing, applying $\boldsymbol{f}$

The purpose of this model is to produce an infiltration matrix, $\boldsymbol{N}$, which defines the weighted closeness of individuals within the social housing population. The model has three components:

- Households, defined by varying numbers of people and sex by age compositions, residing in dwellings.
- Varying numbers and spatially located dwellings in buildings, where a 'building' can be a single dwelling or multi-unit housing building. Multi-unit housing dwellings are positioned in a three-dimensional space by the distance along a corridor, $\boldsymbol{x}$ (as 0,1,2,3 etc.), side of a corridor, $\boldsymbol{y}$ (0 or 1), and level or floor, $\boldsymbol{z}$ (from 0, up to 30 for some buildings).
- A drift function, $\boldsymbol{f}$, that defines the level of second-hand smoke a dwelling (and therefore the individuals in that dwelling) could be exposed to, based on the relative position to other dwellings.

The matrix is modelled with these components as follows:

1. A random building is drawn (weighted by proportion of each building type).
2. Random households are drawn to fill the building dwellings (weighted by proportion of each family type. This proportion includes 'empty' households, i.e., no occupants in a dwelling, to account for <100% occupancy rates).
3. The drift function is applied to each dwelling.

4. The process is repeated 10,000 times, with the infiltration matrix, $\boldsymbol{N}$, produced as the average across all draws. $\boldsymbol{N}$ is input into the PMSLT to calculate the health impact of second-hand smoke exposure alongside direct smoking (described in the next section).

### 5. PMSLT

#### 5.1 PMSLT overview

The proportional multi-state lifetable (PMSLT) is described in detail in Blakely et al. (35) It consists of two components:

1. A main 'all-cause' lifetable, comprising the population living in social housing in Victoria, Australia. All-cause morbidity and mortality rates are projected forward for 20 years based on historic Global Burden of Disease (GBD) study data, then held constant.
2. 31 subsidiary disease lifetables. Each disease is defined by incidence, case fatality, remission, and disability rates, derived from historic GBD disease trends.

The PMSLT, and its link to the Markov and Matrix models, is shown in Supplementary Figure 5.

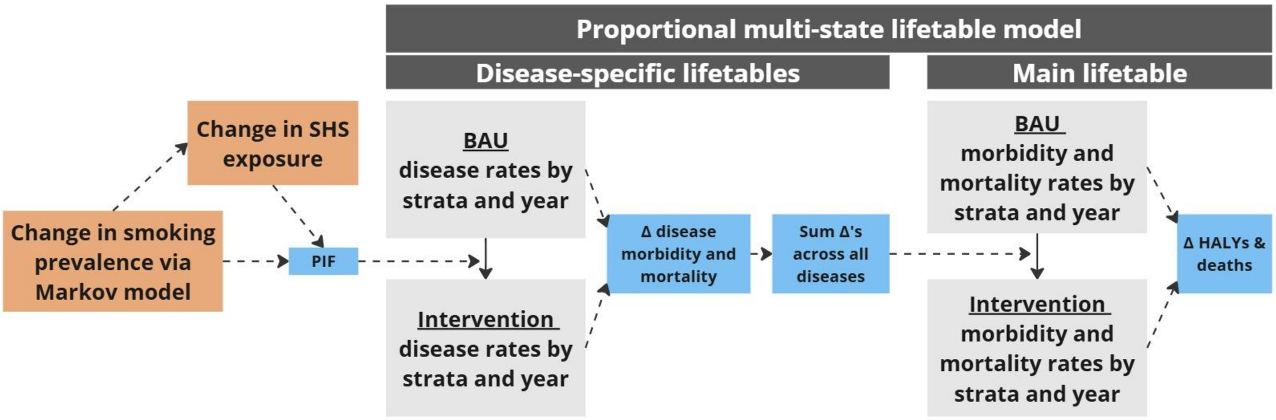


**Supplementary Figure 5.**
Disease rates in each lifetable include incidence, fatality, remission, and disability.
BAU: business-as-usual; HALYs: health-adjusted life years; PIF: potential-impact fraction; SHS: second-hand smoke.

#### 5.2 PMSLT data sources

The inputs for the PMSLT are summarised in Supplementary Table 5.

**Supplementary Table 5. PMSLT data inputs**

| Input | Source | Detail |
|---|---|---|
| Starting population (2023) | Australian Census, 2021, ABS (16)<br>Housing Assistance in Australia report, 2024, AIHW (8) | Social housing population by age, sex, and dwelling type (multi-unit housing vs. separate housing)<br>Census data from 2021 used to estimate population distribution – scaled up to AIHW estimate of 2023 population. |
| Birth projections | ABS (16-19)<br>Housing Assistance in Australia report, 2024, AIHW (8) | [See Supplementary Table 1] |
| Migration projections | Victorian Government (11) | [See Supplementary Table 1] |
| All-cause mortality rate projections | GBD study, IHME (20)<br>Mortality Over Regions and Time books, 2017–2021, AIHW (21) | [See Supplementary Table 1] |
| Smoking and second-hand smoke- related disease parameters (incidence, prevalence, CFR, disability rate, remission) | GBD study, IHME (20)<br>AIHW reports (36-41)<br>NCCI reports (42,43) | Historic GBD incidence, prevalence & CFR values by sex and age for each disease projected forward, and remission rates solved for.<br>Adjusted by SEIFA using AIHW & NCCI data: apply most disadvantaged quintile (SES1) values. |
| Smoking intensity | NDSHS (via Australian Data Archive) (44-51) | Average cigarettes smoked per day (CPD) by sex and age for the most disadvantaged SEIFA quintile estimated using NDSHS data<br>Logistic regression model applied to project CPD, from which pack-years were derived [see section 5.4.1] |
| Rate ratios linking smoking and second-hand smoke exposure to diseases | Dai et al., 2022 (52)<br>Flor et al. (53)<br>National Health Survey, ABS (54,55) | Smoking-linked diseases: lung cancer, low back pain, chronic obstructive pulmonary disease (COPD), ischaemic heart disease (IHD), stroke, diabetes mellitus, oesophageal cancer, pancreatic cancer, Alzheimer's and other dementias, asthma, aortic aneurysm, colorectal cancer, liver cancer, leukaemia, bladder cancer, lower respiratory tract infections (LRTI), larynx cancer, breast cancer, lip and oral cavity cancer, other pharynx cancer, cervical cancer, stomach cancer, kidney cancer, atrial fibrillation and flutter, peripheral artery disease, gallbladder and biliary disease, rheumatoid arthritis, prostate cancer, multiple sclerosis, peptic ulcer disease, and Parkinson's disease.<br>• Risk varies by either CPD or pack-year units.<br><br>Second-hand smoke-linked diseases: ischaemic heart disease, stroke, type 2 diabetes mellitus, lung cancer<br>• Dichotomous values (exposure = yes/no) converted to a continuous linear scale by level of exposure estimated from the matrix model. Assuming that exposure = yes reflects exposure within a typical smoking household – average number of people who smoke per smoking household used to estimate this value. Linear scaling then applied for exposure=0 to maximum value in matrix. No variation by age or sex. |
| Decay in disease risk in years after quitting smoking | Hoogenveen et al. (56) | • Decay function, with decay factor for specific diseases, applied for 20 years post-quitting smoking, followed by a linear decline to RR=1 for each disease at year 30 post-quitting<br>• Decay in disease risk post-exposure to second-hand smoke is estimated indirectly using the population of people who formerly smoked. |

AIHW: Australian Institute of Health and Welfare; NCCI: National Cancer Control Indicators.

##### 5.3 Potential Impact Fraction

Under an intervention scenario, a change in smoking prevalence (and consequently second-hand smoke exposure) alters disease incidence in the PMSLT lifetables via potential impact fractions (PIFs). Direct smoking is linked with all 31 diseases, while second-hand smoke exposure is linked to eight of the diseases.

The resulting changes in specific disease morbidity and mortality are summed in each year and compared to all-cause morbidity and mortality in the BAU model. The model outputs this difference as a change in health-adjusted life years (HALYs) (35), which are calculated in the same way as quality-adjusted life years (QALYs) but use disability weights instead of utility values, given the ready availability of disability weights for each disease (as used by the GBD). (57)

The PIF for those currently or formerly smoking was calculated for each disease in each timestep for every sex by birth cohort, as

$$\boldsymbol{Smoking\ PIF} = \frac{\sum_{j=1}^{n} P_j\ RR_j - \sum_{j=1}^{n} P_j'\ RR_j}{\sum_{j=1}^{n} P_j\ RR_j}$$

where $j$ is the Markov model state ($\boldsymbol{N}, \boldsymbol{S}, \boldsymbol{F_{1-30}}$), $P$ is the proportion of the specific strata in each state $j$ under BAU, compared to the intervention scenario ($\boldsymbol{P'}$).

Second-hand smoke exposure values were calculated within the PMSLT by multiplying the infiltration matrix, $\boldsymbol{N}$, with a vector of smoking prevalence by sex and age in each year. This exposure was then applied to a PIF for each disease in each timestep for every sex by birth cohort, as

$$\boldsymbol{SHS\ PIF} = \frac{\sum_{j=0} P_j RR_j - \sum_{j=0} P_j' RR_j}{\sum_{j=0} P_j RR_j}$$

where $j$ is the continuous second-hand smoke exposure level, $\boldsymbol{P}$ is the population second-hand smoke exposure distribution under BAU compared to the intervention scenario ($\boldsymbol{P'}$).

#### 5.4 Smoking-disease associations

Incidence relative risks (RRs) linking current smoking to each tobacco-related diseases (52) follow dose-response relationships in 'pack-years' (for COPD and cancers, except prostate cancer), or smoking intensity (as cigarette equivalents smoked per day; all other outcomes). For those in the formerly smoked (**F**) states, the RR is scaled down from the value in the final year prior to quitting over 20 years ($\boldsymbol{F_1}$-$\boldsymbol{F_{20}}$) using a decay function, (56) followed up a linear decline to RR=1 by $\boldsymbol{F_{30}}$ (i.e., no increased risk compared to those in the $\boldsymbol{N}$ state 30-years post quitting).

##### *5.4.1 Pack years & smoking intensity*

Smoking intensity is estimated using a log-linear regression model on sex by age by year trends in cigarettes smoked per day (CPD). Pack-years can be calculated from smoking intensity estimates for a given sex by birth cohort, $\boldsymbol{i}$, as

$$\boldsymbol{PY_{i,y}} = \sum_{k=1}^{y} \frac{cig_{k..y}}{20}$$

where $\boldsymbol{k}$=1 is the year of smoking uptake, $y$ is the current year, and $\boldsymbol{cig}$ is the mean cigarettes smoked per day for a given cohort – the 'intensity' component of pack-years. This value is divided by 20, as the usual number of cigarettes in a 'pack'. When interpreting pack-years, $\boldsymbol{PY}$ = 1 can be equivalent to 1 pack per day for a year, or a 10th of a pack (approx. 2 cigarettes) per day for 10 years.

In our model, we specify $\boldsymbol{k}$=1 to be the year in which age = 20; rather than allowing variation in the age at which smoking uptake occurs, we make a simplifying assumption that 20 years is the approximate average age of smoking uptake (given that uptake generally occurs between ages 15-25).

To calculate pack-years for a given birth cohort each year, the smoking intensity from the age of uptake is required. In the base year, 2023, the oldest age is 109 years. Given that age 20 is the age that smoking uptake is assumed to occur for these calculations, smoking intensity needs to be estimated from 1934 for the oldest cohort. Therefore, smoking intensity was both forward and back projected with the regression model, to then derive pack-years.

National Drug Strategy Household Survey (NDSHS) sex by age by SEIFA data from 2001-2022/23 was used as the regression model input, with the model output for the SES1 quintile used (following the process for other model inputs, given a lack of data on smoking intensity for the population living in social housing in Victoria directly). While differences in smoking intensity by sociodemographic group are present in the available NDSHS data, it is unlikely that these differences by SEIFA existed in the early years of the tobacco epidemic. In fact, trends may have been the opposite to what is seen today (consider that today, vaping is more common among more advantaged than disadvantaged groups, the opposite to the current trend for established smoking (58)). Information on smoking intensity by sociodemographic group is not available for the pre-2001 period to specifically inform how these differences by strata changed over time. Applying trends in smoking intensity by remoteness and SEIFA strata for the whole required historic period based on the 2001 to 2022-23 trends would likely result in PYs being overestimated for disadvantaged birth cohorts who accumulate PYs prior to 2001. CPD (and subsequently PYs) were therefore calculated for different time periods as follows:

- **Pre 1970 (required for PY calculation):** CPD predicted by sex and age only. This was done by applying a regression model to NDSHS smoking intensity data (sex by age model below).
- **1971-2000 (required for PY calculation):** CPD predicted by sex and age, then adjusted in each year for differences by SEIFA. This adjustment was done by using CPD predictions by sex, age, remoteness and SEIFA from 2001 (all strata model below), converting these values into ratios of the sex by age totals in 2001, then linearly scaling the ratios by calendar year, moving backward to values of 1 (i.e., no difference SEIFA quintile for a given sex by age group) in 1970. The scaled ratios were applied to the sex by age CPD predictions from 1971-2000, to produce a gradual increase in the relative difference in smoking intensity across SEIFA quintile over this time period.
- **2001-2022 (required for PY calculation):** CPD predicted by sex, age, and SEIFA. This was done by applying a regression model to NDSHS smoking intensity data (all strata model, equation 18 below).
- **2023-2043 (required for CPD and PY calculation):** CPD predicted by sex and age for 2023-2043 (sex by age model below), then adjusted in each year for differences by SEIFA. This adjustment done by using CPD predictions by sex, age, and SEIFA in 2023 (all-strata model below), converting these values into ratios of the sex by age totals in 2023, then applying the ratios to the sex by age CPD predictions in 2023-2043. This assumes that there is no further change in the relative differences in smoking intensity by SEIFA beyond the 2023 level.
- **2044 onward (required for CPD and PY calculation):** CPD values from 2043 (as estimated above) applied in 2044 onward, assuming no further change in smoking intensity by cohort.

The two separate linear regression models used to forward and back predict CPD values are shown below – one containing only sex, age and year variables (sex by age model), and the other additionally including SEIFA (all strata model). For each, the best fit model was selected based on Akaike Information Criterion (AIC), comparing models including age$^2$ and age$^3$ terms and interaction terms between the year, sex, age variables. No interactions terms were included by SEIFA.

$$\boldsymbol{cpd}_{\boldsymbol{sex\ by\ age\ model}} = B_0 + B_1 calendar\ year + B_2 age + B_3 age^2 + B_4 age^3 + B_5 sex \times age.$$

$$\boldsymbol{cpd}_{\boldsymbol{all\ strata\ model}} = B_0 + B_1 calendar\ year + B_2 age + B_3 age^2 + B_4 age^3 + B_5 sex \times age + B_6 seifa.$$

In the above, age (range: 20-70 years) and calendar year are both numerical variables, and SEIFA is categorical. Each regression model was weighted using individual-level sample weights from the NDSHS unit-level files, using the *Survey* package (26) to account for the complex survey design. To account for the statistical imprecision in CPD predictions, the regression models were sampled with replacement 2000 times using bootstrapping to obtain a 95% uncertainty interval (UI) around the mean. This was done in R (version 4.2.2) with the *bootPredict* function. (59)

Ages 71+ were given the same value as 70-year-olds within the same birth cohort. Trends were also truncated by birth year, with the 1931 and 2002 birth years acting as the upper and lower limits (this covers 70-year-olds in 2001-, and 20-year-olds in 2022). Birth years outside of this range were given the same value as the upper or lower limit cohort.

The output of both models were converted to birth cohort values, then the final version of CPD in each year calculated based on the strata rules defined above.

#### 5.5 Second-hand smoke-disease associations

The infiltration matrix, $\boldsymbol{N}$, produced by the matrix model, defines the exposure level of second-hand smoke for every individual in the population in each year. The relevant RR value is assigned by second-hand smoke exposure level, to then be applied to the PIF. Second-hand smoke exposure values were calculated within the PMSLT by multiplying $\boldsymbol{N}$ with a vector of smoking prevalence by sex and age in each year.

##### *5.5.1 Relative risk scaling*

Binary exposure-disease RRs (exposed to second-hand smoke: yes/no) from the GBD (53) were converted to continuous RRs for the PIF equation. Given that exposure in the GBD is based on self-report, living in a smoking household is likely the closest estimate of the amount of second-hand smoke exposure in 'exposed' category. The average number people who smoke per smoking household in Australia is 1.25 (54,55) – so the 'exposed' GBD RR value was mapped to an exposure level of $\boldsymbol{j} = 1.25$. Linear scaling was then used to produce RR values from $\boldsymbol{j} = 0$ (where the RR for each disease is 1) to the maximum level.

#### Supplementary Results

**Supplementary Table 6. Relative difference in health impact (in HALYs gained) from total smoking eradication, under different second-hand smoke parameterisation scenarios**

| Intervention scenario | Alternative BAU scenarios | HALY gain (95% UI) relative to BAU | Percentage change |
|---|---|---|---|
| **Stand-alone housing** | | | |
| **Total smoking eradication [1]** | Main analysis | 12,800 (10,600 to 15,500) | Reference |
| | No SHS exposure | 9,840 (7,730 to 12,400) | 23.1% decrease |
| **Multi-unit housing** | | | |
| **Second-hand smoke eradication** | Main analysis | 2,060 (1,810 to 2,360) | Reference |
| | No smoke drift | 939 (828 to 1,070) | 54.4% decrease |
| | Low smoke drift | 1,910 (1,680 to 2,180) | 7.3% decrease |
| | High smoke drift | 2,170 (1,900 to 2,480) | 5.3% increase |
| **Total smoking eradication [1]** | Main analysis | 6,920 (5,860 to 8,230) | Reference |
| | No SHS exposure | 5,080 (4,050 to 6,320) | 26.6% decrease |
| | No smoke drift | 5,920 (4,720 to 7,260) | 14.5% decrease |
| | Low smoke drift | 6,800 (5,430 to 8,100) | 1.7% decrease |
| | High smoke drift | 7,020 (5,620 to 8,350) | 1.4% increase |

[1]Total smoking eradication results in eradication of second-hand smoke exposure.

Note: no smoke drift in stand-alone housing, hence why smoke drift sensitivity analyses are not included.

BAU: business-as-usual; HALY: health-adjusted life year; SHS: second-hand smoke.

**Supplementary Table 7. Relative difference in health impact (in deaths averted) from total smoking eradication, under different second-hand smoke parameterisation scenarios**

| Intervention scenario | Alternative BAU scenarios | Deaths averted (95% UI) relative to BAU | Percentage change |
|---|---|---|---|
| **Stand-alone housing** | | | |
| **Total smoking eradication [1]** | Main analysis | 892 (757 to 1,040) | Reference |
| | No SHS exposure | 592 (491 to 712) | 33.6% decrease |
| **Multi-unit housing** | | | |
| **Second-hand smoke eradication** | Main analysis | 239 (204 to 282) | Reference |
| | No smoke drift | 92.3 (79.0 to 109) | 61.4% decrease |
| | Low smoke drift | 219 (187 to 259) | 8.4% decrease |
| | High smoke drift | 253 (216 to 299) | 5.9% increase |
| **Total smoking eradication [1]** | Main analysis | 538 (457 to 628) | Reference |
| | No SHS exposure | 337 (280 to 405) | 37.4% decrease |
| | No smoke drift | 408 (345 to 484) | 24.2% decrease |
| | Low smoke drift | 516 (438 to 599) | 4.1% decrease |
| | High smoke drift | 545 (461 to 633) | 1.3% increase |

Total smoking eradication results in eradication of second-hand smoke exposure.

Note: no smoke drift in stand-alone housing, hence why smoke drift sensitivity analyses are not included.

BAU: business-as-usual; SHS: second-hand smoke.

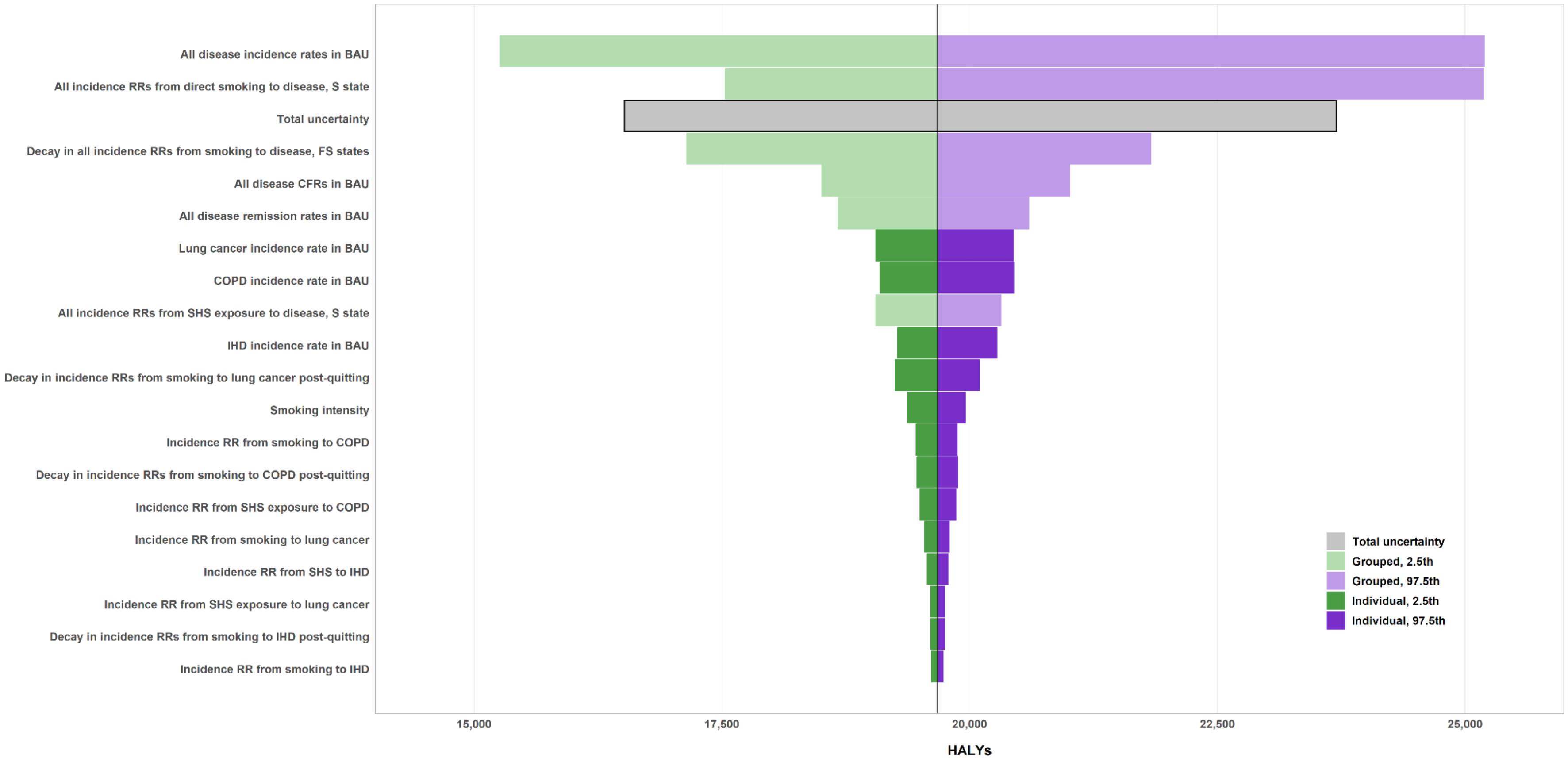


**Supplementary Figure 6. Tornado plot showing change in total health-adjusted life years (HALYs) gained under total smoking eradication scenario in comparison to BAU (2025-2045), with uncertain input parameter groups individually set at the 2.5th and 97.5th percentile values.**

Grouped uncertainty: forces 100% correlation across all diseases for a given factor (e.g., ‘disease incidence rates’ forces incidence rates to be drawn from the 2.5th, then 97.5th, percentile for all 31 diseases in the model) – this results in uncertainty for ‘All disease incidences rates in BAU’ being greater than the total uncertainty, where diseases incidence RRs are not correlated.

Individual uncertainty: three largest contributors to tobacco burden included as individual parameter uncertainty analysis – lung cancer, COPD, and IHD.

IHD: ischemic heart disease; COPD: chronic obstructive pulmonary disease.; RR: relative risk.
